# The Association Between Alcohol Consumption and Anal Human Papillomavirus in the HPV Infection in Men Study

**DOI:** 10.1101/2025.05.23.25328238

**Authors:** Sherissa Mohammed-Ali, Victoria Umutoni, Matthew B. Schabath, Alan G. Nyitray, Bradley Sirak, Eduardo Lazcano-Ponce, Luisa Villa, Anna R. Giuliano, Staci L. Sudenga

## Abstract

**Background:** Human papillomavirus (HPV) can lead to anal cancer in men and women. The aim of this analysis was to assess the association between alcohol consumption and the prevalence and incidence of anal HPV among 1,919 men.

**Methods:** The HPV infection in Men Study recruited men without HIV. Anal specimens were collected at baseline and follow-up visits. Using established methods, prevalence and incidence of HPV was determined. Alcohol consumption was reported as number of days of alcohol consumed per month and categorized into quartiles. Poisson regression was used to determine the association between HPV prevalence and alcohol consumption. A multivariable logistic regression model was used to determine the association between HPV incidence and alcohol consumption. Analyses were stratified by smoking status and sexual orientation.

**Results:** Overall, alcohol consumption was not significantly associated with prevalence or incidence of anal HPV. Current smokers with alcohol consumption of 6+ days per month had a decreased prevalence of high-risk HPV infections (adjusted PR (aPR)=0.43; 95%CI:0.20-0.90) compared to those with 0 days. While former smokers with an alcohol consumption of 3-5 days (aPR=5.29; 95%CI:1.17-25.1) and 6+ days (aPR=5.33; 95%CI:1.15-24.7) had an increased prevalence of high-risk HPV infection compared to 0 days a month.

**Conclusions:** Higher number of days exposed to alcohol among former smokers was significantly associated with anal HPV prevalence. The known inflammatory effect of alcohol may have promoted HPV resulting in higher anal HPV prevalence. Additional research is needed to assess the role of alcohol in anal HPV prevalence among former smokers.

## Introduction

Human papillomavirus (HPV) is a sexually transmitted infection that can lead to anal cancer in both men and women. HPV types 16 and 18 are high-risk HPV (HR-HPV) types that most commonly cause anal cancer.^1^ Anal cancer incidence is highest in the United States and Western Europe.^2^ In 2024, anal cancer accounted for 0.5% of all estimated new cancer cases and has an increasing mortality rate.^3,4^ Anal cancer has surpassed cervical cancer incidence in the US among White women >65 years making it the most common HPV-related cancer in elderly White women.^5^ There is high geographic disparity in anal cancer incidence and mortality in the US with most cases and deaths occurring in the Southeast.^6^

Certain populations are at an increased risk for anal cancer, mainly people with HIV (PWH), men that have sex with men (MSM), women diagnosed with HPV-related gynecological precancerous lesions or cancer, solid organ transplant recipients and patients with autoimmune diseases^7^. Risk factors for anal HPV include a high number of sexual partners, tobacco use, HIV, and sexually transmitted infection (STI) coinfection.^5,8–10^ We have previously shown that there is a higher prevalence of genital HPV infections among men with a higher alcohol intake.^11^ Previous studies have shown that alcohol use and heavy smoking are associated with adverse outcomes for HPV-related oropharyngeal cancer.^12^

High alcohol consumption has been shown to impact both the innate and adaptive immune systems making individuals more susceptible to infections and disease^13^. Alcohol consumption has an effect on proinflammatory cytokine production, impacting the innate immune response.^14^ There is a decrease in the T and B cell response and dendritic cell function following alcohol exposure, indicating that alcohol may impair the adaptive immune response.^15^

There is a limited number of studies evaluating the impact of alcohol consumption on the natural history of anal HPV. The aim of this study was to assess the association between alcohol consumption and the prevalence and incidence of anal HPV in men without HIV from the United States, Brazil and Mexico.

## Methods

### Study Population

The HPV infection in Men (HIM) Study recruited 4123 men from Sao Paulo, Brazil; Cuernavaca, Mexico; and Tampa, Florida between July 2005 and June 2009. Inclusion criteria were HIV-negative men between the ages of 18 to 70 years. The study design for the HIM study has been published previously ^16^. In brief, men completed a pre-enrollment and baseline visit as well as subsequent visits scheduled 6 months apart for up to ten years. Participants had a physical exam and completed a Risk Factor Questionnaire which details sexual history and sociodemographic characteristics.

The study team obtained written informed consent from all subjects. Study procedures were authorized by the Institutional Review Boards at the University of South Florida (Tampa, FL), the Ludwig Institute for Cancer Research, the Centro de Referencia e Treinamento em Doencas Sexualmente Transmissiveis e AIDS (Sao Paulo, Brazil), and the Instituto Nacional de Salud Publica (Cuernavaca, Mexico).

### Anal Specimen collection for HPV detection

Participants that consented to anal specimen collection had a specimen collected at each visit. Details of anal specimen collection has been published elsewhere^10^. Anal specimens underwent DNA extraction (Qiagen Media Kit) followed by PCR analysis and HPV genotyping (Roche Linear Array). Samples showing positive results for beta-globin or any HPV genotype were analyzed. HPV genotyping tested for 37 HPV types which were categorized as having any HPV, high-risk (HR-HPV) or low-risk (LR-HPV) HPV. HR-HPV includes types 16, 18, 31, 33, 35, 39, 45,51, 52, 56, 58, 59, and 68. LR-HPV includes types 6, 11, 26, 40, 42, 53, 54, 55, 61, 62, 64, 66, 67, 69, 70, 71, 72, 73,81, 82, 82 subtype IS39, 83, 84, and 89.^17^

### Statistical Analysis

Participants with missing alcohol data (n=190) were excluded from the analyses. At baseline, participants self-reported their lifetime number of sexual partners and sex was defined as oral, anal, or vaginal sex. Sexual orientation was assigned based on sex of sexual partners reported at baseline and we categorized participants as men who have sex with men regardless of having sex with women (MSM) or men who have sex with women only (MSW). Participants who did not report on sexual partners were included as never had sex for sexual orientation. Lifetime number of male or female sexual partners were each categorized as 0-1, 2-9, 10-49, 50+ or missing. Tobacco smoking status was categorized as current smoker, former smoker or never.

Participants were asked several alcohol consumption questions in the baseline survey. Participants reported the number of days drinking alcohol per month and the average number of alcohol drinks consumed on days when they drank. Alcohol consumption number of days drinking per month was categorized into quartiles (Q1: 0 days Q2: 1-2 days, Q3: 3-5 days, Q4: 6+ days) given that there is not a recommend cut point for days of alcohol consumption in the literature. Number of alcohol drinks per day was also categorized into quartiles but was not significantly associated with anal HPV prevalence or incidence in any of the analyses.

### HPV Prevalence

HIM Study participants with an anal HPV genotyping results at baseline were included in the analysis (n=1919). The prevalence of any HPV, HR-HPV and LR-HPV was assessed. Prevalence of any HPV is defined as having any of the 37 HPV genotypes present at the anal canal. HR-HPV and LR-HPV occurs when one or more of those specific subsets of HPV genotypes are present. To examine the association between HPV prevalence and alcohol consumption, prevalence ratio (PR) and 95% confidence intervals were examined using a Poisson Regression with robust variance estimation. The outcome variables for the models includes the following: (i) Any HPV vs. No HPV, (ii) HR-HPV vs. no HR-HPV (iii) LR-HPV vs. no LR-HPV.

The multivariable model was adjusted based on potential confounders that were chosen *a priori*. PRs of the overall cohort was adjusted for age, country, and lifetime male and female sexual partners, and smoking status. The prevalence ratios were stratified by sexual orientation and smoking status.^8,18^ PRs of the MSM model were adjusted for country, age, and smoking status and male and female sexual partners. PRs of the MSW model were adjusted for country, age, smoking status and female sexual partners. In the current, former and never models, the PRs were adjusted for are age, country, and male and female sexual partners. In a Poisson regression analysis, the effect of interaction between smoking status and alcohol consumption on prevalent anal HPV infections was examined.

### HPV Incidence

Participants with anal HPV specimens collected at subsequent visits were included in the analysis of incident anal HPV infections(n=1427). An incident anal HPV infection was defined as the presence of new type specific infection that was not present at baseline. Incidence of any HPV, a HR-HPV or LR-HPV genotype correspond to having at least one those HPV types being newly detected during a follow-up visit. To determine the association between incident HPV infections and days of alcohol consumption, odds ratios (OR) and 95% confidence intervals (CI) were calculated using a multivariable logistic regression. The odds ratios were presented as an overall cohort and then stratified by sexual orientation and smoking status. The models were adjusted for age, country, lifetime male and female sexual partners and smoking status. Odds ratios were stratified by sexual orientation and smoking status.

### Data availability

Data that support this study are only available upon requests. They are not publicly accessible due to the delicate nature of the data presented in this study. All data pertinent to this study are incorporated in the article or uploaded as Supplementary Material. Study dataset with deidentified participant data and protocol can be requested from.

## Results

### Baseline Characteristics

Among study participants in this analysis (n=1919), 45% are between the ages of 18 to 30, 39% are between 31 to 44, and 15% are between the ages of 45 to 70 (**Table 1**). About 45% of participants that drank 0 days a month were Brazilian; comparably, 53% of participants that drank 6+ days a month were from the US. Within the study, 23% of participants were current smokers, 20% were former smokers and 58% had never smoked. Among participants, 20% were MSM and 69% were MSW.

**Table 1.** Baseline demographic and sexual characteristics by days of alcohol consumption per month in quartiles.

| Characteristic | Overall<br>(n=1919) | Days of alcohol consumption per month |  |  |  |
| --- | --- | --- | --- | --- | --- |
|  |  | 0 (Q1)<br>(n=447) | 1-2 (Q2)<br>(n=513) | 3-5 (Q3)<br>(n=474) | 6+ (Q4)<br>(n=485) |
| **Age** |  |  |  |  |  |
| 18-30 | 872 (45.4) | 158 (35.3) | 234 (45.6) | 238 (50.2) | 242 (49.9) |
| 31-44 | 754 (39.3) | 198 (44.3) | 216 (42.1) | 182 (38.4) | 158 (32.6) |
| 45-70 | 293 (15.3) | 91 (20.4) | 63 (12.3) | 54 (11.4) | 85 (17.5) |
| **Country of Residence** |  |  |  |  |  |
| Brazil | 669 (34.9) | 202 (45.2) | 154 (30.0) | 173 (36.5) | 140 (28.9) |
| Mexico | 633 (33.0) | 135 (30.2) | 247 (48.1) | 163 (34.4) | 88 (18.1) |
| US | 617 (32.1) | 110 (24.6) | 112 (21.8) | 138 (29.1) | 257 (53.0) |
| **Smoking Status** |  |  |  |  |  |
| Current | 433 (22.6) | 65 (14.5) | 115 (22.5) | 115 (24.3) | 138 (28.4) |
| Former | 379 (19.8) | 100 (22.4) | 80 (15.6) | 84 (17.7) | 115 (23.7) |
| Never | 1106 (57.7) | 282 (63.1) | 317 (61.9) | 275 (58.0) | 232 (47.8) |
| Missing | 1 (0.1) | 0 (0) | 1 (0.2) | 0 (0) | 0 (0) |
| **Race** |  |  |  |  |  |
| Asian/PI | 39 (2.03) | 9 (2.01) | 8 (1.56) | 12 (2.53) | 10 (2.06) |
| Black | 284 (14.8) | 80 (17.9) | 73 (14.2) | 62 (13.08) | 69 (14.23) |
| Other | 658 (34.3) | 147 (32.9) | 252 (49.1) | 165 (34.8) | 94 (19.4) |
| White | 896 (46.7) | 202 (45.2) | 165 (32.2) | 225 (47.5) | 304 (62.7) |
| Refused | 42 (2.19) | 9 (2.01) | 15 (2.92) | 10 (2.11) | 8 (1.65) |
| **Ethnicity** |  |  |  |  |  |
| Hispanic | 891 (46.9) | 202 (45.6) | 313 (61.2) | 229 (49.1) | 147 (30.6) |
| Non-Hispanic | 1010 (53.1) | 241 (54.4) | 198 (38.7) | 237 (50.9) | 334 (69.4) |
| Missing | 18 (0.9) | 4 (0.9) | 2 (0.4) | 8 (1.7) | 4 (0.8) |
| **Marital Status** |  |  |  |  |  |
| Cohabiting | 220 (11.5) | 46 (10.3) | 70 (13.7) | 61 (12.9) | 43 (8.88) |
| Divorced/Separated/ Widowed | 180 (9.41) | 47 (10.6) | 33 (6.46) | 44 (9.32) | 56 (11.6) |
| Married | 669 (35.0) | 186 (41.8) | 214 (41.9) | 141 (29.9) | 128 (26.4) |
| Single | 843 (44.1) | 166 (37.3) | 194 (38.0) | 226 (47.9) | 257 (53.1) |
| Missing | 7 (0.4) | 2 (0.5) | 2 (0.4) | 2 (0.4) | 1 (0.2) |
| **Sexual Orientation** |  |  |  |  |  |
| MSM | 390 (20.3) | 99 (22.1) | 86 (16.8) | 102 (21.5) | 103 (21.2) |
| MSW | 1322 (68.9) | 282 (63.1) | 360 (70.2) | 335 (70.7) | 345 (71.1) |
| Refused | 207 (10.8) | 66 (14.8) | 67 (13.1) | 37 (7.81) | 37 (7.63) |
| **Lifetime Male Sexual Partners** |  |  |  |  |  |
| None to one | 1589 (82.8) | 361 (80.8) | 444 (86.6) | 383 (80.8) | 401 (82.7) |
| Two to nine | 158 (8.23) | 41 (9.17) | 27 (5.26) | 45 (9.49) | 45 (9.28) |
| 10 to 49 | 89 (4.64) | 22 (4.92) | 19 (3.70) | 25 (5.27) | 23 (4.74) |
| 50+ | 46 (2.40) | 10 (2.24) | 11 (2.14) | 13 (2.74) | 12 (2.47) |
| Missing | 37 (1.93) | 13 (2.91) | 12 (2.34) | 8 (1.69) | 4 (0.82) |
| **Lifetime Female Sexual Partners** |  |  |  |  |  |
| None to one | 368 (19.2) | 110 (24.6) | 100 (19.5) | 87 (18.3) | 71 (14.6) |
| Two to nine | 765 (39.9) | 174 (38.9) | 224 (43.7) | 208 (43.9) | 159 (32.8) |
| 10 to 49 | 593 (30.9) | 112 (25.1) | 140 (27.3) | 144 (30.4) | 197 (40.6) |
| 50+ | 111 (5.78) | 23 (5.15) | 27 (5.26) | 21 (4.43) | 40 (8.25) |
| Missing | 82 (4.27) | 28 (6.26) | 22 (4.29) | 14 (2.95) | 18 (3.71) |

Abbreviations: Men that have sex with men (MSM); men that have sex with women (MSW)
Sexual Orientation Refused: Participants who did not report on sexual partners were included as refused for sexual orientation

### HPV Prevalence by Days of Alcohol Consumption

The prevalence of any HPV genotype ranged from 16.9% (Q4) to 20.3% (Q3) for days of alcohol consumption (**Table 2**). The prevalence of a HR-HPV genotype ranged from 8.8% (Q2) to 12.2% (Q3). When stratified by smoking status, the prevalence of HR-HPV genotype among current smokers was 23.1% for Q1 and 11.6% for Q4. The prevalence of HR-HPV genotype among former smokers was 2.0% for Q1 and 10.4% for Q4. Prevalence of HR-HPV among MSM ranged from 25.2% (Q1) to 33.0% (Q4) and among MSW ranged from 5.0% (Q2) to 7.0% (Q3).

**Table 2.** Prevalence and 95% CI of HPV genotypes by alcohol consumption and smoking status.

|  | **Any HPV**<br>**PR (95% CI)** | **HR-HPV**<br>**PR (95% CI)** | **LR-HPV**<br>**PR (95% CI)** |
| --- | --- | --- | --- |
| **Overall** |  |  |  |
| 0 (Q1) | 18.3 (14.9-22.2) | 11.2 (8.42-14.5) | 13.9 (10.8-17.4) |
| 1-2 (Q2) | 17.5 (14.4-21.1) | 8.77 (6.47-11.6) | 12.7 (9.92-15.9) |
| 3-5 (Q3) | 20.3 (16.7-24.2) | 12.2 (9.42-15.6) | 14.6 (11.5-18.1) |
| 6+ (Q4) | 16.9 (13.7-20.6) | 11.3 (8.66-14.5) | 11.1 (8.48-14.3) |
| **Current Smokers** |  |  |  |
| 0 (Q1) | 30.1 (20.0-43.4) | 23.1 (13.5-35.2) | 20.0 (11.1-31.8) |
| 1-2 (Q2) | 23.5 (16.1-32.3) | 12.2 (6.82-19.6) | 17.4 (11.0-25.6) |
| 3-5 (Q3) | 20.9 (13.9-29.4) | 8.70 (4.25-15.4) | 19.1 (12.4-27.5) |
| 6+ (Q4) | 19.6 (13.3-27.2) | 11.6 (6.77-18.1) | 14.5 (9.08-21.5) |
| **Former Smokers** |  |  |  |
| 0 (Q1) | 11.0 (6.0-19.0) | 2.0 (0.2-7.0) | 11.0 (5.6-18.8) |
| 1-2 (Q2) | 16.0 (8.0-26.0) | 6.25 (2.06-14.0) | 15.0 (8.0-24.7) |
| 3-5 (Q3) | 15.0 (9.0-25.0) | 10.7 (5.02-19.4) | 9.52 (4.2-17.9) |
| 6+ (Q4) | 18.3 (11.7-26.6) | 10.4 (5.51-17.5) | 13.9 (8.2-21.6) |
| **Never Smokers** |  |  |  |
| 0 (Q1) | 18.1 (13.8-23.1) | 11.7 (8.19-16.0) | 13.0 (9.0-18.0) |
| 1-2 (Q2) | 15.8 (11.9-20.3) | 8.20 (5.43-11.8) | 10.0 (7.0-14.0) |
| 3-5 (Q3) | 21.4 (16.8-26.8) | 14.2 (10.3-18.9) | 14.2 (10.3-18.9) |
| 6+ (Q4) | 14.7 (10.4-19.9) | 11.6 (7.81-16.5) | 8.0 (5.0-12.0) |
| **MSM** |  |  |  |
| 0 (Q1) | 41.4 (31.6-51.8) | 25.2 (17.1-35.0) | 32.3 (23.3-42.5) |
| 1-2 (Q2) | 43.0 (32.4-54.1) | 24.4 (15.8-34.9) | 30.2 (20.8-41.1) |
| 3-5 (Q3) | 49.0 (39.0-59.1) | 31.4 (22.6-41.3) | 40.2 (30.6-50.4) |
| 6+ (Q4) | 40.8 (31.2-50.9) | 33.0 (24.1-43.0) | 28.2 (19.7-37.9) |
| **MSW** |  |  |  |
| 0 (Q1) | 9.9 (6.7-14.0) | 6.0 (3.6-9.5) | 7.4 (4.7-11.2) |
| 1-2 (Q2) | 12.5 (9.3-16.4) | 5.0 (3.0-7.8) | 9.2 (6.4-12.6) |
| 3-5 (Q3) | 12.2 (8.9-16.2) | 7.5 (4.9-10.8) | 6.9 (4.4-10.1) |
| 6+ (Q4) | 12.5 (9.3-16.4) | 5.5 (3.3-8.5) | 6.7 (4.3-9.8) |
| **Never Had Sex\*** |  |  |  |
| 0 (Q1) | 11.9 (6.06-22.1) | 9.0 (4.06-18.6) | 8.95 (4.1-18.6) |
| 1-2 (Q2) | 11.9 (6.06-22.1) | 2.7 (0.4-17.0) | 13.5 (5.7-28.7) |
| 3-5 (Q3) | 10.8 (4.09-25.6) | 5.4 (1.3-19.3) | 5.40 (1.3-19.3) |
| 6+ (Q4) | 19.7 (11.8-31.1) | 12.1 (6.2-22.4) | 13.6 (7.2-24.2) |

HR-HPV types: 16, 18, 31, 33, 35, 39, 45, 51, 52, 56, 58, 59, and 68
LR-HPV types: 6, 11, 26, 40, 42, 53, 54, 55, 61, 62, 64, 66, 67, 69, 70, 71, 72, 73, 81, 82, 82 subtype IS39, 83, 84, and 89
Any HPV types: HR-HPV and LR-HPV described above
Abbreviations: Men that have sex with men (MSM); men that have sex with women (MSW)
Sexual Orientation Never Had Sex\*: Participants who did not report on sexual partners were included as never had sex for sexual orientation

### Relationship between Days of Alcohol Consumption and HPV Prevalence

Overall, there was no significant difference in anal HPV prevalence by days of alcohol consumption after adjusting for age, country, smoking and sexual orientation (**Table 3**). In a Poisson regression model, an overall test of interaction between smoking status and alcohol consumption on prevalent high-risk anal HPV infections was statistically significant (p = 0.015). Models were stratified by smoking status and sexual orientation. There was no significant association between alcohol consumption and anal HPV prevalence when stratified by sexual orientation (**Table 3**). After stratifying by smoking status, there was no significant association between alcohol consumption and anal HPV infection among never smokers. Current smokers had a decreased prevalence of HR-HPV infections with an alcohol consumption of 3-5 days a month (Q3, aPR=0.28; 95%CI: 0.12-0.66) and 6+ days a month (Q4, aPR=0.43; 95%CI: 0.20-0.90) Former smokers had a higher prevalence of HR-HPV infections with an alcohol consumption of 3-5 days a month (Q3, aPR=5.54; 95%CI: 1.14-26.9) and 6+ days a month (Q4, aPR=5.42; 95%CI: 1.11-26.4) compared to those with 0 days of alcohol consumption (Q1).

**Table 3.**
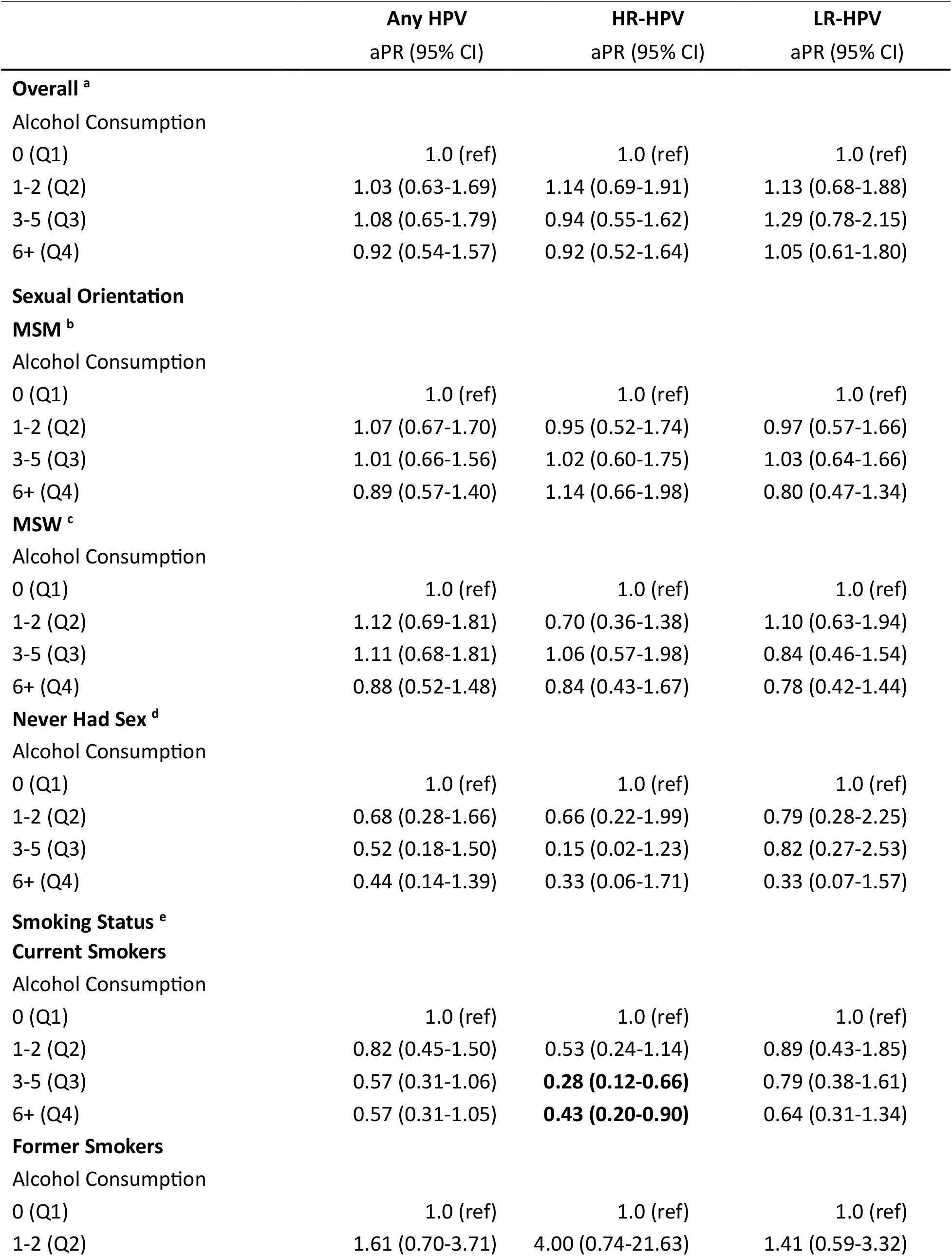
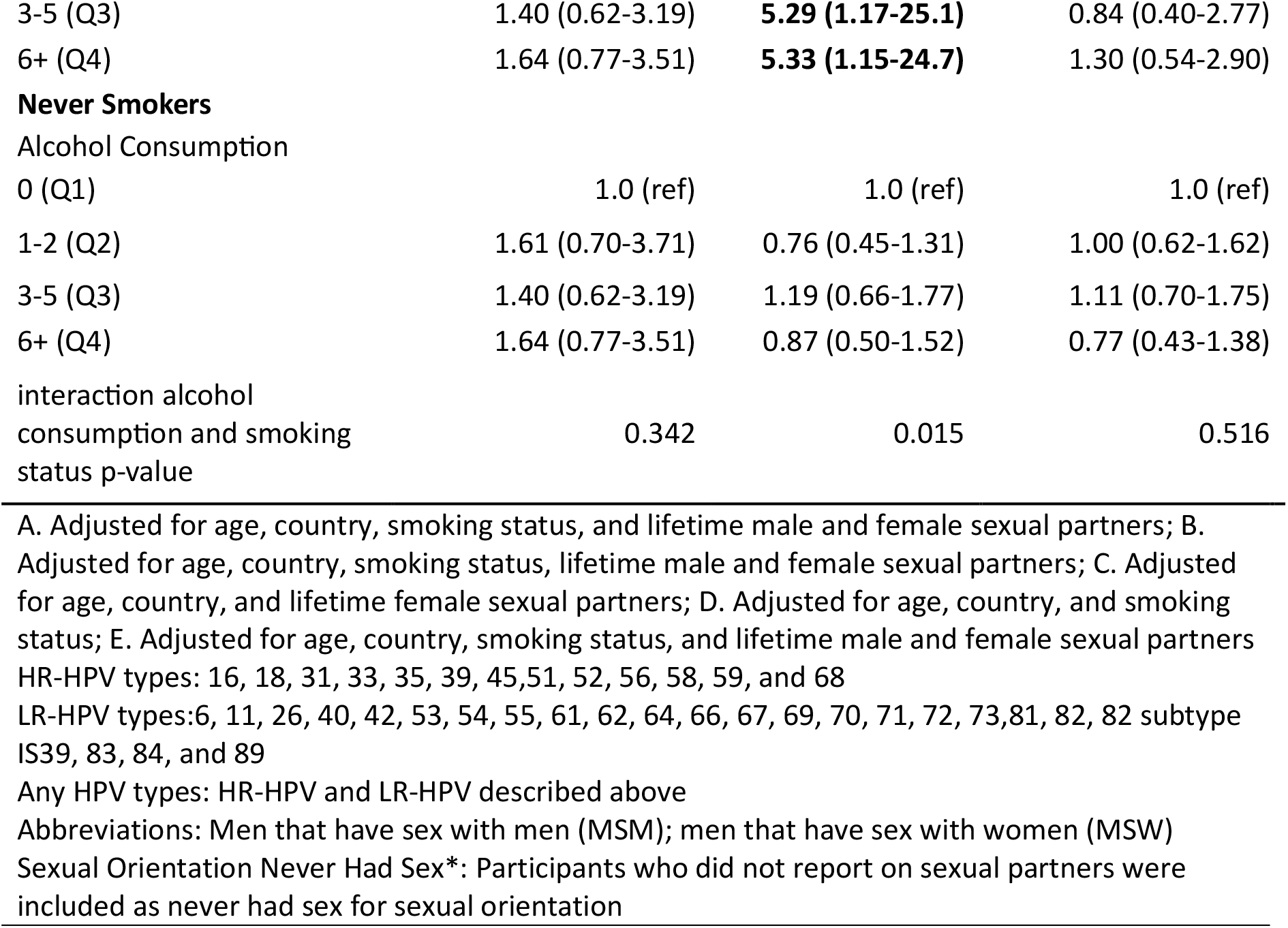
Adjusted Prevalence Ratio (aPRs) for association between alcohol intake and anal HPV prevalence by sexual orientation and smoking status.

### Relationship between Days of Alcohol Consumption and HPV Incidence

Overall, there was no significant association between days of alcohol consumption and incident anal HPV infections after adjusting for age, country, smoking status, and lifetime male and female sexual partners (**Table 4**). Additionally, when stratified by sexual orientation and smoking there was no significant association between days of alcohol consumption and incident anal HPV infections (**Table 4**).

**Table 4.**
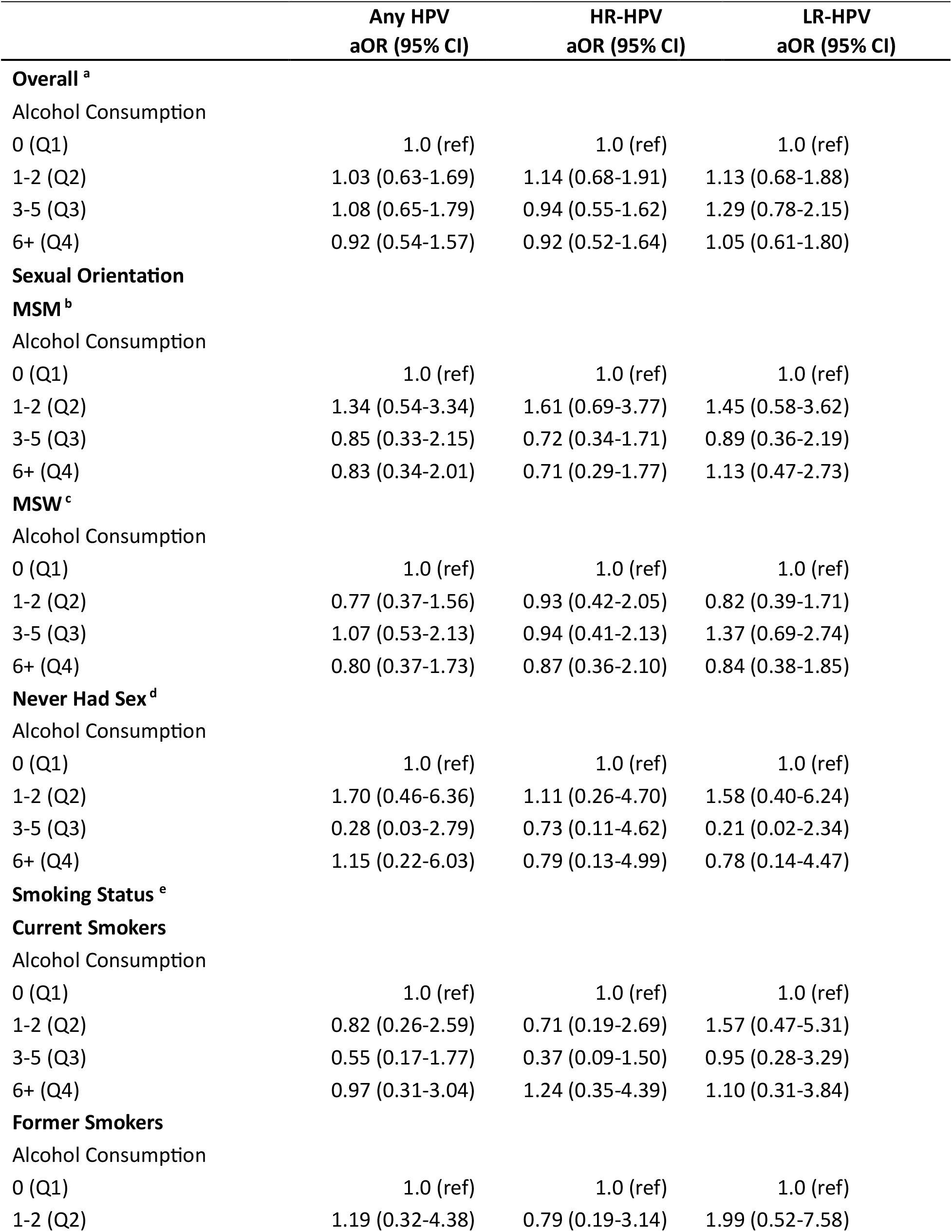
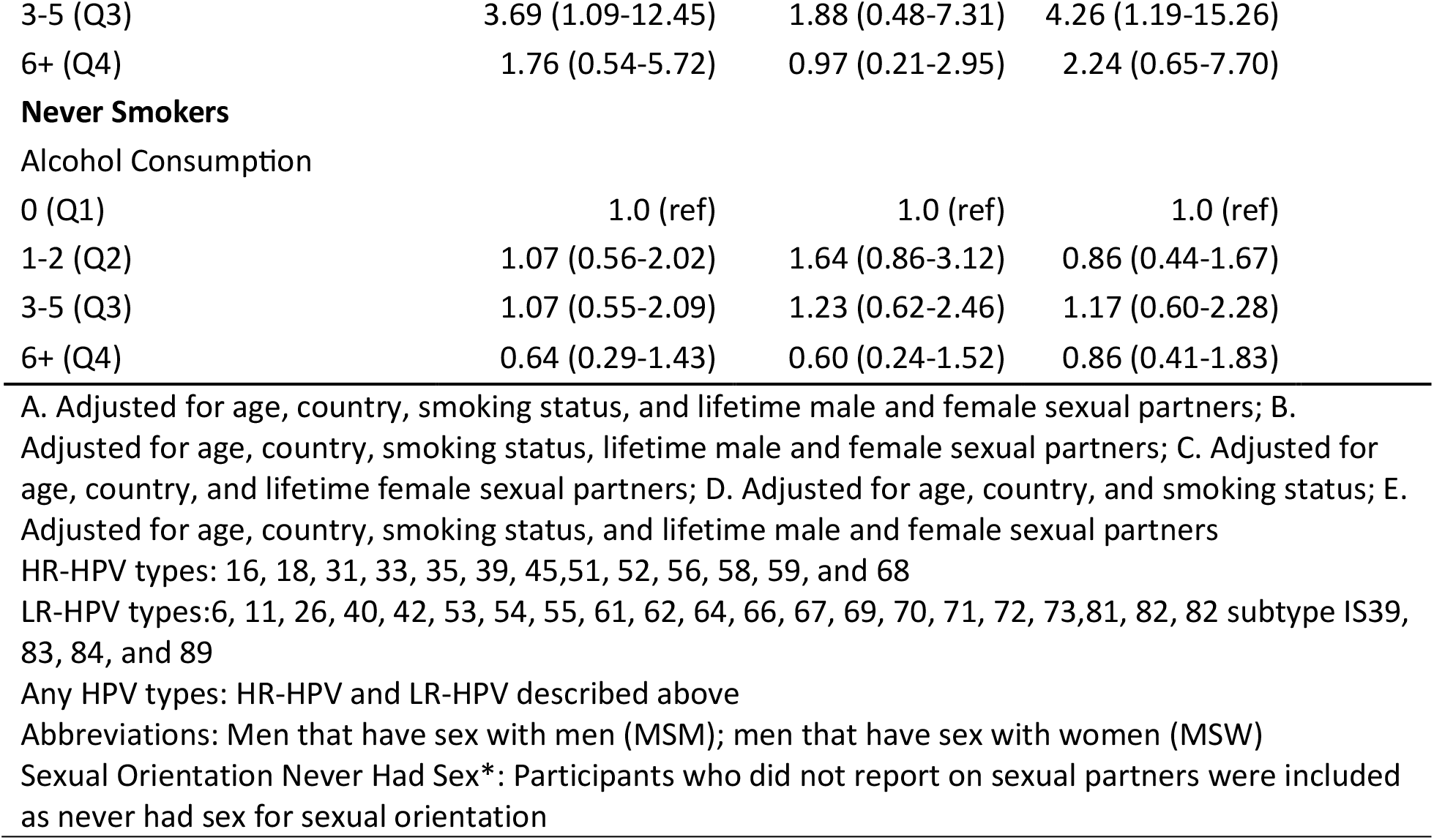
Adjusted Odds Ratio for the association between alcohol intake and anal HPV incidence by sexual orientation and smoking status.

## Discussion

Among a HIV-negative population of men from the US, Brazil and Mexico, the association between days of alcohol consumption and prevalent and incident anal HPV infections were examined. Overall, there was no statistically significant association between prevalent or incident anal HPV infections and days of alcohol consumption per month. However, when stratified by smoking status, there was a significant association between alcohol consumption and prevalent high-risk anal HPV infections among current and former smokers. Former smokers had a significantly increased prevalence of high-risk anal HPV infections with high number of days of alcohol consumption. Current smokers had a significantly decreased prevalence of high-risk anal HPV infections with a high number of days of alcohol consumption. There was a significant interaction between smoking status and alcohol consumption on high-risk anal HPV prevalence. Smoking status modifies the effect of alcohol consumption on high-risk anal HPV infection prevalence.

The HIM Study has previously examined the role of smoking status as a risk factor for prevalent, incident and persistent anal HPV infections^10^. In determining the association between anal HPV infections and smoking status, there was significantly higher prevalence and incidence of anal HPV infections among current smokers when compared to participants that had never smoked. There was no significant difference in anal HPV infections between former smokers and participants that had never smoked^10^. Using data from the HIM Study, *Schabath et al.*, identified a significant association between higher alcohol consumption (defined by number of drinks per month) and prevalent genital HPV infections. However, after stratifying by smoking status, there was no significant association between alcohol consumption and prevalent genital HPV infections.^11^ These findings contrast with the results of this study that indicates a decreased prevalence of high-risk anal HPV infections with increasing days of alcohol consumption among current smokers. The examination of the association between alcohol consumption and genital HPV was restricted to the US and quantified alcohol consumption as number of drinks, causing differences in the results between the genital and anal HPV studies.

Previous studies have examined the combined effects of smoking status and alcohol consumption on HPV status and cancer-related outcomes.^18^ Current, but not former, smoking significantly increased the risk of high-grade squamous intraepithelial lesions (HSIL) at the cervix in high-risk HPV-positive women. Alcohol consumption showed no association with HSIL risk.^19^ Thakral et al observed both smoking and alcohol consumption independently increased the risk of HPV-positive head and neck squamous cell carcinoma.^20^ Smoking status and alcohol consumption may interact to impact HPV-related outcomes, particularly in head and neck cancers. Studies indicate combined exposure to tobacco and alcohol exacerbate HPV infection, increase cancer risk and worsen prognosis.

Higher alcohol consumption greatly impacts both innate and adaptive immune processes. Afshar et al observed binge alcohol consumption caused an early, transient pro-inflammatory response followed by an anti-inflammatory state, affecting circulating monocytes and cytokine production.^21^ Beech et al observed that alcohol dependence is associated with altered expression of genes in cytokine signaling pathways.^22^ Days of alcohol consumption in this study were associated with increased HPV prevalence among former smokers but HPV prevalence was not associated with the amount of alcohol consumed. It is possible that former smokers are replacing the urge to smoke with alcohol consumption ^23^ and this is leading to other changes in behavior among these former smokers that may also be playing a role in higher HPV prevalence. More research is needed to fully understand the role of smoking and alcohol consumption in anal HPV natural history.

This study has many strengths including a large sample size with participants from the US, Mexico and Brazil. The study considers any HPV, HR-HPV and LR-HPV and assesses multiple known factors of HPV status such as sexual orientation and smoking status. Few previous studies of anal HPV have been conducted in an HIV-negative population or among MSW. The HIM Study questionnaire included several questions related to alcohol consumption, which allowed for an in-depth investigation into the role of alcohol consumption in the anal HPV natural history. However, the current study has several limitations that should be considered. The HIM Study is not a population-based study, but the demographics such as race, ethnicity, education, and age of the men included at each clinical site are similar to the underlying population of men aged 18-70 years in their respective communities ^24,25^. We conducted multivariable models to adjust for lifetime sexual partners but not does not capture the different types of sexual intercourse that could be associated with risk. Additionally, lifetime sexual partners may not reflect current sexual behaviors at the time of anal canal sampling which may explain the lower anal HPV prevalence among MSM participant in our study compared to others. Stratified analyses by smoking status may have been limited in power given the limited precision in some of the estimates evident by the wide confidence intervals.

To conclude, increasing days of alcohol consumption was associated with high-risk anal HPV prevalence among former smokers. Among current smokers, those that did not drink had a higher prevalence of high-risk anal HPV. Both smoking and alcohol consumption are pro-inflammatory and may increase HPV persistence. Among former smokers, there may be residual pro-inflammatory state due to alcohol consumption which increases susceptibility to anal HR-HPV infections. Further studies on the combined effects of smoking status and alcohol consumption on anal HPV infection is needed to understand the role of alcohol consumption on anal HPV duration and progression to disease.

## Acknowledgements

This research was supported in part by research funding from Merck Sharp & Dohme Corp. The opinions expressed in this paper are those of the authors and do not necessarily represent those of Merck Sharp & Dohme Corp.

We thank the HIM Study teams and participants in the United States (Moffitt Cancer Center, Tampa), Brazil (Centro de Referência e Treinamento em DST/AIDS, Fundação Faculdade de Medicina Instituto do Câncer do Estado de São Paulo, Ludwig Institute for Cancer Research, São Paulo), and Mexico (Instituto Mexicano del Seguro Social, Instituto Nacional de Salud Pública, Cuernavaca).

